# The clinical characteristics and mortal causes analysis of COVID-19 death patients

**DOI:** 10.1101/2020.04.12.20062380

**Authors:** Ao-Xiang Guo, Jia-Jia Cui, Qian-Ying OuYang, Li He, Cheng-Xian Guo, Ji-Ye Yin

## Abstract

**Purpose:** Currently, COVID-19 is causing a large number of deaths globally. However, few researches focused on the clinical features of death patients. This study conducted a retrospective analysis of clinical characteristics and mortal causes in Chinese COVID-19 death patients.

**Patients and methods:** The clinical characteristics of death patients were collected from publicized by local health authorities in China. Expressions of virus targets in human organs were obtained from GTEx database.

**Results:** 159 patients from 24 provinces in China were recruited in our study, including 26 young patients under 60 and 133 aged 60 or older. The median age was 71 years, which indicated that most death patients were elderly. More male patients died of COVID-19 than females (1.65 fold). Hypertension was the most common coexisting disorder and respiratory failure was the most common direct cause of death. Fever (71.19%) and cough (55.08%) were the predominant presenting symptoms. There was one asymptomatic patient. In addition, by comparing young and old patients, heart disease was identified as an important risk factor for death in the aged patients. ACE2 and TMPRSS2 were the targets of SARS-CoV-2, we analyzed their expression in different organs. TMPRSS2 and ACE2 had a high expression in the organs which had corresponding clinical features in death patients.

**Conclusion:** Male, age and heart disease were the main risk factors of death. Beside, asymptomatic patients with serious coexisting disorders may also die of SARS-CoV-2. Thus, more attention should be paid to the old patients with heart disease and asymptomatic patients in the treatment.

## 1. Introduction

COVID-19 was firstly reported in Wuhan, Hubei Province, China at the end of last year [1, 2]. Currently, it is breaking out globally. As of April 11, COVID-19 has cause over 1.7 million infections and over 100 thousand deaths around the world. In China, more than 3,300 patients infected with COVID-19 died. The clinical manifestations of COVID-19 infectors were already summarized [1, 3]. However, few studies focused on the clinical characteristics of death patients. It is important to summarize these data to find out the potential risk factors that can help identify patients with poor prognosis at an early stage. Furthermore, clear understanding of the direct causes of death is very important to reduce death. However, what are the causes of death has never been answered in previous studies.

It was reported that membrane protein angiotensin converting enzyme II (ACE2) and transmembrane serine protease 2 (TMPRSS2) were the targets of SARS-CoV-2 in human [4-6]. Therefore, we supposed that the expression of ACE2 and TMPRSS2 in human tissues could be used to explain the clinical characteristics of COVID-19 patients, including coexisting disorders, direct causes of death and initial symptoms.

In this study, we did a comprehensive evaluation for death patients in China. Besides, the expression of SARS-CoV-2 targets in different organs were analyzed. We aimed to summarize the clinical characteristics of death patients, and thus to provide information for reducing death and developing effective treatment.

## 2. Methods

### 2.1 Data collection

159 anonymous dead patients infected with COVID-19 publicized by local health authorities were recruited. All patients were diagnosed with COVID-19 based on the National Health Commission’s Protocol of Diagnosing & Treating COVID-19 from 28 December, 2019 to 4 March, 2020. Clinical features including gender, age, geographic location, clinical symptoms, coexisting disorders and days from illness to death were collected. The GTEx database (https://www.gtexportal.org) was used for analyzing the expression of ACE2 and TMPRSS2. The study protocol was approved by Institutional Review Board (AAHRPP-accredited) of the Third Xiangya Hospital of Central South University.

### 2.2 Data processing

We performed descriptive statistical analysis of ratios and percentages. We used the number of deaths divided by the number of confirmed cases to calculate daily mortality of COVID-19. Statistical analyses in this study were performed with use of SPSS 17.0 software (IBM, NY, USA). Categorical variables were present as numbers and percentages. Continuous variables were present as median and interquartile range if they were not normally distributed. We compared proportions for categorical variables by using the χ2 test. A two-sided P value less than 0.05 was considered statistically significant.

## 3. Results

### 3.1 Basic clinical characteristics

Based on our collected data, the mortality in different provinces in China is different. In order to randomly select representative samples, we collected the data of mortality in both Hubei which is the most-affected province and other 23 provinces since 25 January, 2020 to 20 March, 2020. As expected, the mortality of COVID-19 in Hubei was much higher than that in other provinces (Figure1). Then we recruited 69 patients from Hubei province and 90 patients from other provinces in mainland China. Of all 159 death patients, 99 were male and 60 were female. The number of males was 1.65 fold than that of female patients. The median age of 159 patients was 71 (interquartile range 64-80) years. The median days from onset of the disease to death was 15 (interquartile range 10-20) days (Table. 1). Thus, male and age were important risk factors for death.

**Figure 1.**
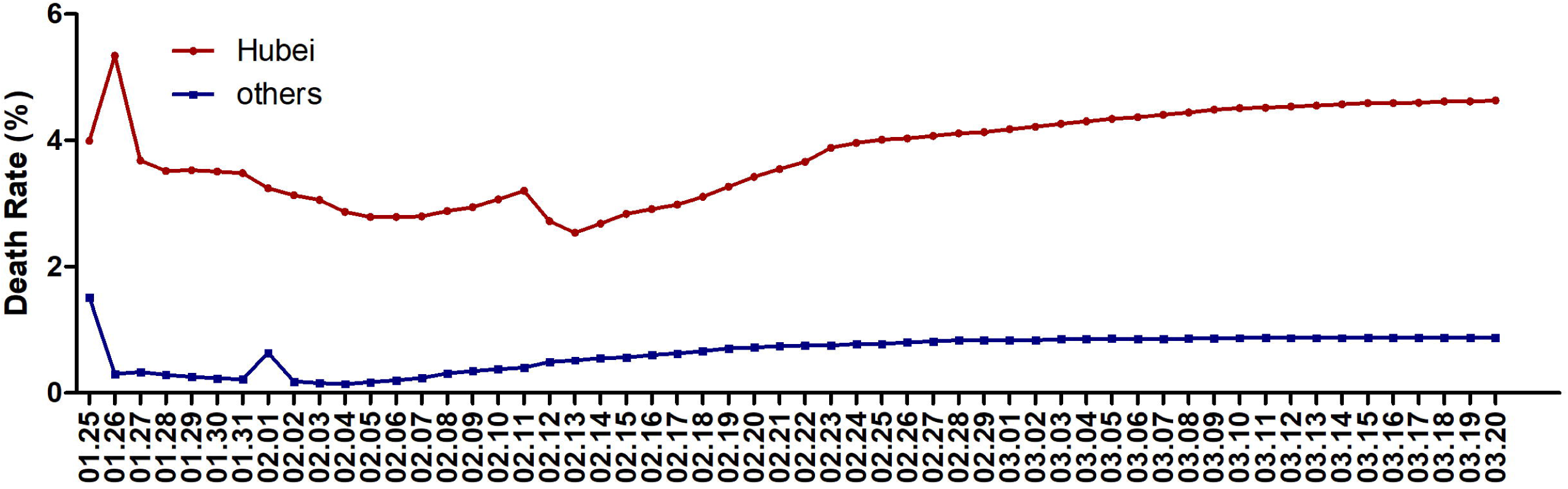
Daily mortality in Hubei and other provinces from January 25 to March 20, 2020.

### 3.2 Coexisting disorders, initial symptom, and treatment

The presence or absence of coexisting disorders may be closely related to the survival of patients. Among 116 patients with information about coexisting disorders, nearly all (99.14%) of them had coexisting disorders. The most common coexisting disorder was hypertension (56.90%), followed by heart disease (38.79%), diabetes (32.76%) and cerebral infarction (14.66%). In addition, there were a few patients with liver diseases, bronchitis, renal diseases, tumor and obesity. These common coexisting disorders focused on cardiovascular diseases such as hypertension and heart disease, which were often occurred in the old people.

After infecting with SARS-CoV-2, most patients had initial symptoms. The most common symptom was fever (71.19%), followed by cough (55.08%), exhaustion (14.41%) and chest tightness (10.17%). Besides, the clinical symptoms occupying a lower proportion should also be noted, including dyspnea, diarrhea, muscle soreness, chills, headache and palpitate and vomiting. It should be noteworthy that 1 death didn’t have initial symptom. These analyses indicated that the initial symptoms were mainly occurred in the lungs, including cough, chest tightness and dyspnea. However, other symptoms such as palpitate and diarrhea should also be paid attention.

As for clinical treatment, most patients received anti-infection therapy (65.26%), antiviral therapy (55.79%) and mechanical ventilation (47.37%). Oxygen treatment and tracheal cannula was account for 38.95% and 32.63%, respectively. Lower percentage of patients received traditional Chinese medicine treatment, anti-inflammatory therapy and dispelling phlegm. Besides, extracorporeal membrane oxygenation (ECMO) and transfusion of plasma donated by recovered patients were also actively used for treatment.

In summary, most death patients had coexisting disorders of cardiovascular diseases such as hypertension. Apart from fever, initial symptoms of death patients were most occurred in the lung. Anti-infection therapy and antiviral therapy were most frequently used for treatment. Our results also proved that coexisting disorders of hypertension and heart disease and initial symptoms of dyspnea were significantly higher in death patients, which was consistent with the one previous study [7].

### 3.3 Direct causes of death

The direct reasons leading to patients death were various. There were 121 patients with information about their direct causes of death. Of them, a majority of patients died of respiratory disorder including respiration failure (47.39%) and acute respiratory distress syndrome (ARDS) (5.79%), which was followed by multiple organs failure (28.93%), heart disease (14.05%) and circulatory failure (13.22%). Cardiac arrest (11.57%) and shock (10.74%) were also important causes of death. Besides, liver injury/failure, renal injury/failure, cerebrovascular accident and disseminated intravascular coagulation (DIC) also contribute to death. Overall, the top three common causes including respiration failure, multiple organs failure and heart disease need special attention.

### 3.4 Comparison between young and old patients

Based on the results above, age was an important risk factor for death. Most (83.65%) death were over 60 years old. However, it is still unknown whether there is a difference between the clinical characteristics of young and old death patients. Therefore, we did a comprehensive evaluation of young and old death patients using these data (Table 1).

**Table 1.** Clinical features COVID-19 death patients

| Clinical features | Total (n=159) | Young (n=26) | Old (n=133) |
| --- | --- | --- | --- |
| **Age groups** |  |  |  |
| 25-59 years | 9/159, 5.66% | 26/159, 16.35% | / |
| 60-94 years | 133/159, 83.64% | / | 133/159, 83.65% |
| **Sex** |  |  |  |
| Male | 99/159, 62.26% | 19/26, 73.08% | 80/133, 60.15% |
| Female | 60/159, 37.74% | 7/26, 26.92% | 53/133, 39.85% |
| **Locations** |  |  |  |
| Hubei | 69/159, 42.86% | 13/26, 50.00% | 56/133, 42.11% |
| Other 23 provinces | 90/159, 56.60% | 13/26, 50.00% | 77/133, 57.89% |
| **Initial symptom** |  |  |  |
| Unknown | 41/159, 25.79% | 4/26, 15.38% | 37/133, 27.82% |
| Fever | 84/118, 71.19% | 19/22, 86.36% | 65/96, 67.71% |
| Cough | 65/118, 55.08% | 15/22, 68.18% | 50/96, 52.08% |
| Exhaustion | 17/118, 14.41% | 3/22, 13.64% | 14/96, 14.58% |
| Chest tightness | 12/118, 10.17% | 1/22, 4.55% | 11/96, 11.46% |
| Dyspnea | 9/118, 7.63% | 0/22, 0.00% | 9/96, 9.38% |
| Diarrhea | 7/118, 5.93% | 1/22, 4.55% | 6/96, 6.25% |
| Muscle soreness | 4/118, 3.39% | 1/22, 4.55% | 3/96, 3.13% |
| Chills | 3/118, 2.54% | 0/22, 0.00% | 3/96, 3.13% |
| Headache | 3/118, 2.54% | 1/22, 4.55% | 2/96, 2.08% |
| Palpitate | 2/118, 1.69% | 0/22, 0.00% | 2/96, 2.08% |
| Vomiting | 2/118, 1.69% | 0/22, 0.00% | 2/96, 2.08% |
| No symptom | 1/118, 0.85% | 0/22, 0.00% | 1/96, 1.04% |
| **Clinical treatment** |  |  |  |
| Unknown | 64/159, 40.25% | 11/26, 42.31% | 53/133, 39.85% |
| Anti-infection therapy | 62/95, 65.26% | 12/15, 80.00% | 50/80, 62.50% |
| Antiviral therapy | 53/95, 55.79% | 7/15, 46.67% | 46/80, 57.50% |
| Mechanical ventilation | 45/95, 47.37% | 5/15, 33.33% | 40/80, 50.00% |
| Oxygen treatment | 37/95, 38.95% | 7/15, 46.67% | 30/80, 37.50% |
| Tracheal cannula | 31/95, 32.63% | 5/15, 33.33% | 26/80, 32.50% |
| Chinese medicine treatment | 13/95, 13.68% | 2/15, 13.33% | 11/80, 13.75% |
| Anti-inflammatory therapy | 12/95, 12.63% | 4/15, 26.67% | 8/80, 10.00% |
| Dispelling phlegm | 11/95, 11.58% | 1/15, 6.67% | 10/80, 12.50% |
| ECMO | 10/95, 10.53% | 4/15, 26.67% | 6/80, 7.50% |
| Adrenaline | 4/95, 4.21% | 1/15, 6.67% | 3/80, 3.75% |
| Dopamine | 2/95, 2.11% | 0/15, 0.00% | 2/80, 2.50% |
| Plasma donated by recovered patients | 2/95, 2.11% | 1/15, 6.67% | 1/80, 1.25% |
| **Coexisting disorder** |  |  |  |
| unknown | 43/159, 27.04% | 11/26, 42.31% | 32/133, 24.06% |
| Hypertension | 66/116, 56.90% | 7/15, 46.67% | 59/101, 58.42% |
| Heart disease | 45/116, 38.79% | 2/15, 13.33% | 43/101, 42.57% |
| Diabetes | 38/116, 32.76% | 6/15, 40.00% | 32/101, 31.68% |
| Cerebral infarction | 17/116, 14.66% | 2/15, 13.33% | 15/101, 14.58% |
| Liver diseases | 10/116, 8.62% | 1/15, 6.67% | 9/101, 8.91% |
| Bronchitis | 10/116, 8.62% | 0/15, 0.00% | 10/101, 9.90% |
| Renal insufficiency | 9/116, 7.76% | 0/15, 0.00% | 9/101, 8.91% |
| Tumor | 6/116, 5.17% | 0/15, 0.00% | 6/101, 5.94% |
| Obesity | 2/116, 1.72% | 2/15, 13.33% | 0/101, 0.00% |
| No Comorbidity | 1/116, 0.86% | 0/15, 0.00% | 1/101, 0.99% |
| **Direct cause of death** |  |  |  |
| Unknown | 38/159, 23.90% | 8/26, 30.77% | 30/133, 22.56% |
| Respiratory failure | 58/121, 47.93% | 6/18, 33.33% | 52/103, 50.49% |
| ARDS | 7/121, 5.79% | 0/18, 0.00% | 7/103, 6.80% |
| Multiple organ failure | 35/121, 28.93% | 6/18, 33.33% | 29/103, 28.16% |
| Heart diseases | 17/121, 14.05% | 1/18, 5.56% | 16/103, 15.53% |
| Circulatory failure | 16/121, 13.22% | 2/18, 11.11% | 14/103, 13.59% |
| Cardiac arrest | 14/121, 11.57% | 4/18, 22.22% | 10/103, 9.71% |
| Shock | 13/121, 10.74% | 1/18, 5.56% | 12/103, 11.65% |
| Sepsis | 8/121, 6.11% | 2/18, 11.11% | 6/103, 5.83% |
| Liver injury / failure | 4/121, 3.31% | 0/18, 0.00% | 4/103, 3.88% |
| Renal injury / failure | 4/121, 3.31% | 0/18, 0.00% | 4/103, 3.88% |
| Cerebrovascular accident | 4/121, 3.31% | 1/18, 5.56% | 3/103, 2.91% |
| DIC | 3/121, 2.48% | 0/18, 0.00% | 3/103, 2.91% |
| **Days from illness to death** |  |  |  |
| Unknown | 42/159, 26.42% | 4/26, 15.38% | 38/133, 28.57% |
| Medians (IQR) | 15 (10-20) | 16.5 (10-24) | 15 (10-19) |

IQR= interquartile range

According to WHO standard, all patients were divided into 2 groups by age: young patients (<60 years) and old patients (≥60 years). As for coexisting disorders, more old patients had a history of heart disease than young patients (42.57% vs 13.33%) (P=0.03). Besides, bronchitis, renal insufficiency and tumor were only occurred in a few old patients and obesity was only found in a few young patients. There was no statistical difference between young and old patients regarding the common direct cause of death and initial symptoms. However, the kinds of coexisting disorders, direct cause of death and initial symptom were more various in old deaths. It is remarkable that there was one asymptomatic patient with coexisting disorder of serious heart failure and renal failure in the old patients group. (Table 1)

In summary, old patients had more kinds of coexisting disorders. Heart disease was one main coexisting disorders and was more frequently occurred in old patients. Therefore, heart disease may be an important risk factor of old patients and should be paid more attention during the treatment.

### 3.5 Virus targets expression

ACE2 and TMPRSS2 were the targets of SARS-CoV-2 in human. It was reasonable to speculate that organs with more expression of virus targets may be more vulnerable to virus attack. Thus, the expression of ACE2 and TMPRSS2 in all major human organs was analyzed.

As Figure 2 showed, TMPRSS2 was expressed highly in lung which may lead to attack of SARS-CoV-2 in lung and cause respiratory failure, ARDS, initial symptoms of cough, dyspnea, and chest tightness. The high expression of TMPRSS2 in liver and kidney may contribute to injury or failure in liver and kidney. TMPRSS2 was also highly in stomach, which may be associated with the initial symptoms of vomiting. While that ACE2 was highly expressed in heart may be related with the attack of SARS-CoV-2 in heart, which may lead to the initial symptoms of palpitate and chest tightness in some patients, and the direct causes of death including circulatory failure, heart disease and cardiac arrest. ACE2 also had a high expression in adipose, which may be related with obesity. Besides, both TMPRSS2 and ACE2 had a high expression in transverse colon and small intestine-terminal ileum, which may be associated with the initial symptom of diarrhea. (Figure 2)

**Figure 2.**
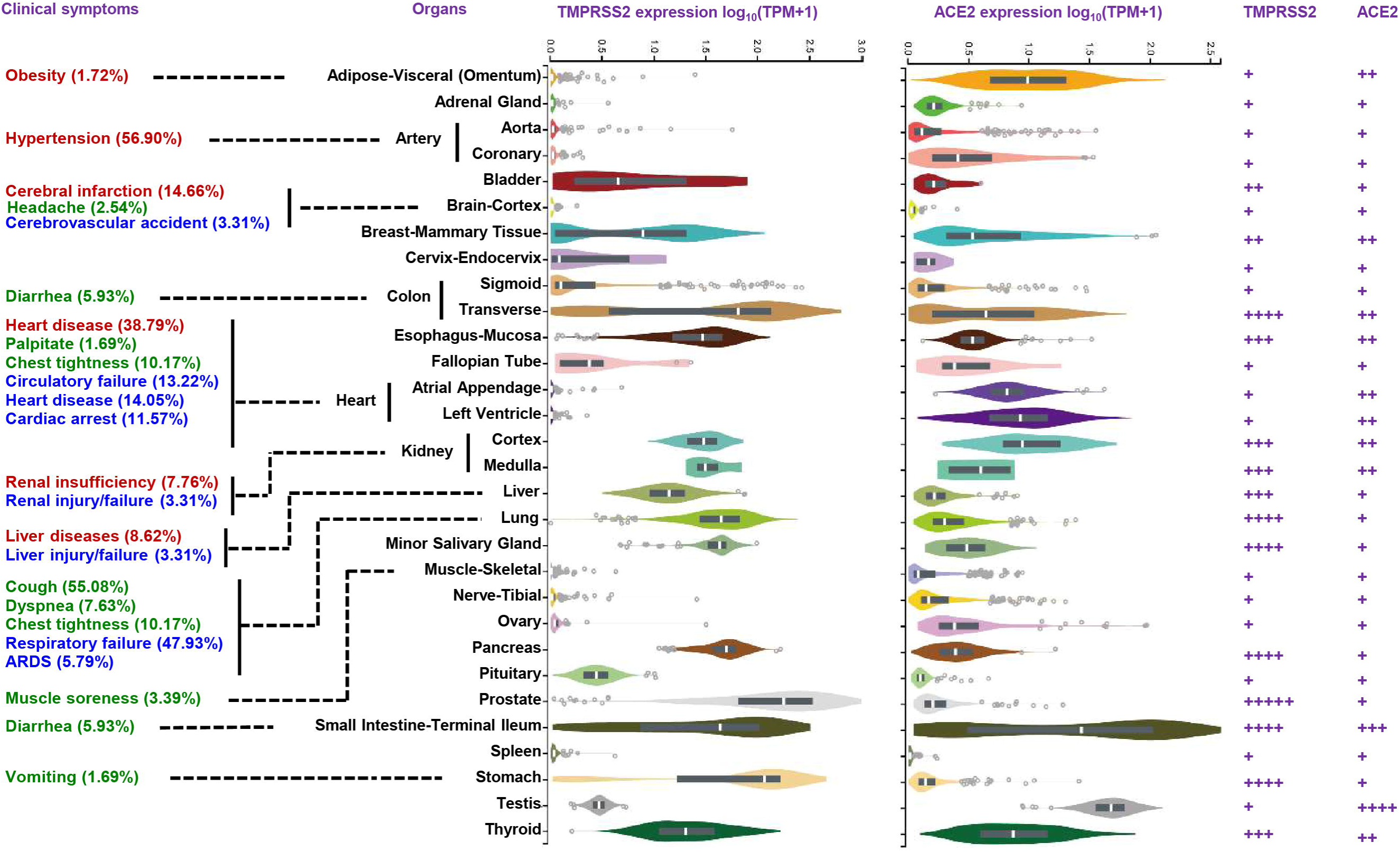
Comprehensive analysis of clinical features and the expression of TMPRSS2 and ACE2 in different organs. Clinical features included initial symptoms (green), coexisting disorders (red) and direct causes of death (blue). The quantified expression grades of TMPRSS2 and ACE2 were shown by the number of “+” in the right.

Taken these results together, TMPRSS2 or ACE2 was highly expressed in the organs with clinical characteristics of dead patients. The expression of the SARS-CoV-2 targets in these important organs such as lung, heart, liver and kidney may help to explain the clinical characteristics of death patients. However, the two targets were also highly expressed in some other organs without obvious clinical symptoms such as pancreas, thyroid, breast, prostate and thyroid. These organs may also be injured but didn’t cause obvious clinical symptoms, which should also be paid attention.

## 4. Discussion

In this study, we found the mortality of COVID-19 in Hubei was significantly higher than that in other provinces of China. Male and age were important risk factors of death. Though the clinical features of young and old people were different, heart disease was one of the most common coexisting disorders and causes of death. In addition, heart disease was more common in the old patients, which may indicate that it was an important risk factor for old patients. It is remarkable that one old dead patient was asymptomatic infector, which suggested that asymptomatic infection in old patients with coexisting disorders may tend to death and should require special attention. Besides, in order to explain the clinical characteristics of death, we analyzed the expression of SARS-CoV-2 targets and found the organs with high expression of TMPRSS2 or ACE2 also had corresponding clinical symptoms in death patients. These findings suggest that older male patients, especially these with heart disease need extra attention to reduce mortality.

The higher mortality in Hubei province may be caused by more patients and less medical resources in the early stages of COVID-19 outbreak. Of all 159 death around China, males was 1.65 fold than female, which indicated that male tend to be easier to death than females. This result was consistent with previous studies which reported that the number of males was more than females among COVID-19 patients [3]. The median age of death patients was 71 years and only 16.35% of patients were under 60 years. Previous study reported the median age of 161 recovered patients was 51 years [7]. This indicates that age is another risk factor of death. Heart disease was the second common coexisting disorders and more old patients had coexisting disorder of heart disease. The high expression of ACE2 in heart make patients with heart disease more weaker and even died. Therefore, we should pay special attention to old patients with heart disease and take measures to protect their hearts.

In order to explain the clinical features of death patients, we combined the clinical features with the expression of ACE2 and TMPRSS2. TMPRSS2 had a high expression in more organs such as lung, liver and kidney, but had a low expression in heart. ACE2 was highly expressed in heart. Therefore, the key target controlling the invasion of SARS-CoV-2 in different organs may be different. And TMPRSS2 seems had a wider expression and may be more important for patients’ infection. Besides, a recent study has reported that a TMPRSS2 inhibitor approved for clinical use could block SARS-CoV-2 from entering cells and might constitute a treatment option [4]. These results provide information for developing effective treatment for COVID-19.

It is interesting to compare the direct causes of death between SARS and COVID-19. The SARS-CoV caused a total of 8,422 infections and 919 deaths in 2003 Different from SARS with mortality of 11% [8]. SARS-CoV-2 had a lower mortality but stronger infectivity. Nearly all SARS infectors had initial symptoms, mainly fever [9]. However, a few SARS-CoV-2 infectors didn’t have any initial symptoms, which emphasized again the importance of asymptomatic patients. Most patients who have died from SARS had co-existing disorders such as acute renal impairment (6.7%) and proteinuria (84.6%) [10, 11]. While more SARS-CoV-2 death patients had cardiovascular diseases such as hypertension (56.90%), which may indicate that the targeted organs of the two virus may be different. In addition, injury or failure of respiratory system and different organs including lung, heart, liver and kidney were important causes of SARS-CoV-2 death. Though the mortality of COVID-19 is lower, it is more complicated.

Recently, more and more asymptomatic infection were found. It is very difficult to aware their infection. However, they also showed strong virus transmission power [12, 13]. Our study found one asymptomatic infection patient. It’s an 80 year old man with coexisting disorders of severe heart failure and renal failure. He died 2 days after diagnosis. This suggested that asymptomatic patients with severe underlying diseases may also die. However, there was only one asymptomatic infection patient in our study. The clinical characteristics of these patients may need to be further explored. This remind us that asymptomatic patients not only have strong transmission ability, but also may be seriously ill and even died.

Several previous studies reported the clinical features of death [7, 14, 15]. However, all of them only include deaths in Wuhan city. This study firstly recruits the largest sample size of death cases from 24 provinces of China and link the direct cause of death with the expression of virus target. Although there have been studies comparing the clinical characteristics of dead and recovered patients and found dead patients were older than recovered patients [7], the risk factors of old deaths had not been identified. We firstly compared the difference of clinical difference between young and old patients and found heart disease was an important risk factor for death of old patients. There were some limitations in our study. The most important one is limited sample size, especially for the stratified analysis. There is a large gap between the sample size of young and old patients, which may cause some findings to be missed. In addition, we may ignore some important information because of the heterogeneity of anonymous COVID-19 data promulgated by local health authorities. We hope that these results can provide valuable information for effective cure and reducing death of COVID-19.

## Data Availability

The data of all anonymous dead patients infected with COVID-19 was publicized by local health authorities in China.

## Acknowledgment

This work was supported by the National Natural Science Foundation of China (81773823, 81573463, 81974511), National Science and Technology Major Project of China (2017ZX09304014, 2019ZX09201-002-006, 2020ZX09201010).

